# An ensemble method associates prepregnancy BMI and maternal ethnicity with key cord blood metabolomic changes in a multi-ethnic cohort from Hawaii

**DOI:** 10.1101/2025.08.14.25333702

**Authors:** Leyang Tao, Bowei Li, Yuheng Du, Shenyu Hung, Lana Garmire

**Author notes:** these authors contributed equally to this work (Leyang Tao, Bowei Li, Yuheng Du).

## Abstract

Maternal obesity poses significant risks to fetal health, influencing metabolomic profiles in newborn cord blood. Despite the growing application of metabolomics, limited research has explored how BMI-associated metabolite alterations may vary across different ethnic groups. We analyzed metabolomic data from a multi-ethnic cohort of 87 participants, including Native Hawaiian and Pacific Islander (NHPI) individuals. We used an ensemble machine learning model with a meta-learner to predict cord blood metabolomic changes associated with maternal BMI, the continuous obesity metric. The meta-learner integrated linear and nonlinear approaches and achieved significantly enhanced performance compared to the baseline linear regression model. In cord blood samples, glycine, serine, and threonine metabolism are activated by maternal obesity, while fatty acid biosynthesis and biosynthesis of unsaturated fatty acids are repressed. Some metabolites associated with these pathways show ethnicity-specific patterns. Compared to Asians and caucasians, 1,5-anhydrosorbitol, glycine, L-threonine show a unique increase from normal to obese maternally associated groups in NHPI, while PC(O-44:6) is significantly decreased in NHPI. The finding reveals the impact of maternal obesity on offspring health, and calls on future research to investigate the maternal and newborn health in underrepresented populations, such as NHPI.

## Introduction

Maternal obesity has been an increasingly alarming health issue. Centers for Disease Control and Prevention (CDC) reported an increasing trend of prepregnancy obesity in the US from 26.1% in 2016 to 29.0% in 2019 ^1^. Maternal obesity may lead to obstetric complications such as cephalopelvic disproportion, preeclampsia^2^. Further, it can induce the risks of transgenerational diseases and complications, including obesity, coronary heart disease, and other neurocognitive and behavioral outcomes.^3–6^ Body Mass Index (BMI) value is generally considered a surrogate metric for obesity. A BMI more than 25 is considered overweight and greater than or equal to 30 is considered obese according to the World Health Organization (WHO)^7^.

Previous studies showed that metabolomic changes in cord blood were significantly associated with the maternal BMI category ^8–11^. Differences were observed in the umbilical cord blood metabolites collected from overweight or obese mothers and mothers of normal pre-pregnancy weight, in metabolites such as corticosterone, 11-deoxycortisol, cortisol, and testosterone, as well as metabolic pathways such as steroid hormone biosynthesis and neuroactive ligand-receptor interactions ^8^. Additionally, ethnicity may affect these metabolomic patterns as well. However, reports addressing underrepresented ethnic groups, particularly the Native Hawaiann and Pacific Islanders (NHPI) are rare. We previously reported that metabolites such as galactonic acid and 2-hydroxy-3-methylbutyric acid were downregulated in the cord blood of newborns born to obese mothers compared to those born to normal-weight mothers^9^. In particular, 2-hydroxy-3-methylbutyric acid appeared to be higher in Native Hawaiians and other Pacific Islanders (NHPI). Such metabolomic changes in lipids, fatty acids (FA), and amino acids (AA) associated with maternal obesity have been linked to aberrant fetal development, increasing the risk of childhood obesity and other growth-related abnormalities.^3,11^

However, this report had several limitations. The first was to categorize the mothers into case and control classes for simplicity, without accounting for the granularity of the obesity metric as reflected by the continuous nature of BMI values. Another potential weakness of the study was relying on the results of elastic-net to identify maternal obesity-associated metabolites, which may be biased due to the limited samples in training the models. Opposite to individual machine-learning methods, the ensemble method that integrates the results of multiple machine-learning models is an alternative approach which has proven particularly effective due to its ability to enhance performance and reduce overfitting ^12^. Traditional ensemble methods such as bagging ^13^, boosting ^14^, and stacking ^15^, along with other models have demonstrated superior flexibility, accuracy, and adaptability across various domains, including genomics and proteomics ^16^.

To address these limitations, we re-analyzed the previous study to associate the metabolomic changes in newborn cord blood with maternal pre-pregnancy obesity represented as the continuous BMI values which preserve the granularity of data, as compared to dichotomized measures which potentially led to information loss and misinterpretation of the relationship between the metabolomic predictors and obesity responders.^17^ Additionally, we applied a stacking ensemble machine learning method to enhance prediction performance and interpretability. Furthermore, we investigated ethnic differences in metabolomic associations, with a focus on NHPI populations, who remain significantly underrepresented in medical research. This integrated approach allows for a deeper understanding of the interplay between BMI, metabolomics, and ethnicity.

## Methods

### Data Acquisition

The data were collected from Kapiolani Medical Center for Women and Children in Honolulu, Hawaii (2015-2018), as previously reported ^9^. The metabolomic raw data files as well as processed data are available at the Metabolomics Workbench repository (https://www.metabolomicsworkbench.org/, study ID ST001114). The study was approved by the Western IRB board (WIRB Protocol #20151223). All participants involved in this study provided written informed consent before the collection of cord blood samples. The cohort consisted of full-term pregnant women scheduled for elective cesarean section (≥37 weeks of gestation). Pre-pregnancy BMI was used to define cases (BMI ≥30.0) and controls (BMI 18.5–25.0) previously. Subjects with pregnancy complications, infections, multiple gestations, or other known confounders were excluded according to criteria detailed in the original study.

### Metabolomic data preprocessing

The metabolite data were collected from three distinct batches, with Batch 1 (N=36), Batch 2 (N=21), and Batch 3 (N=30). Missing values in the dataset were imputed using the “impute”^18^ package in R, employing a KNN-based imputation method. After the imputation, variance stabilizing normalization (VSN) was performed to reduce the variance within each metabolite across samples.^19^ Combat function from the “sva” package was applied to the normalized data^20,21^ to adjust for potential batch effects. To assess potential confounders affecting metabolomic variation, we conducted the analysis of variance (ANOVA) test using the “car” ^22^ R package across demographic and technical variables (**Table1**; **Supplementary Figure1**).

### Feature selection and model performance based on the Ensemble model

An ensemble model was built to predict the continuous BMI value based on the metabolite level and confounders included in the model (Equation 2.1).

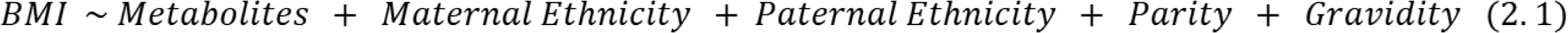

The ensemble framework comprised three base models: Random forest^23^, XGBoost^24^, and Elastic-net^25^. A meta-model of linear regression combined the predictions of these three base learners to optimize the ultimate performance. Random forest and Elastic net were implemented with the “sklearn” package^26^, while the XGBoost model was accessed through the “XGBoost” package in Python^24^ with default settings. The root mean square error (RMSE) and Pearson Correlation Coefficient (PCC) were obtained from 5-fold cross-validation with 10 repeats to indicate the average performance of the model. The feature importance was explained by averaging the absolute Shapley value^27^ over 50 iterations.

### Pathway Analysis

Metabolite enrichment analysis was performed using the “Lilikoi2” package^28^. Metabolites were mapped to Kyoto Encyclopedia of Genes and Genomes (KEGG) pathways stored in the Lilikoi database^29^. Among the 185 metabolites selected in the preprocessing, 170 of them (91.9%) mapped to 27 distinct metabolic pathways, each pathway containing at least three metabolite hits in KEGG dataset ^28^. Pathway deregulation scores (PDS) were computed using the “Pathifier” package^30^, focusing on pathways containing at least three metabolites. The PDS indicates how a pathway is deregulated from normal status, with a range from 0 to 1, higher number indicates more deviation. Kendall’s tau correlation test^31^ was applied to determine the association between the pathway deregulation score and continuous BMI value. To correct for multiple hypothesis testing, p-values from Kendall’s tau correlation tests were adjusted using the Benjamini–Hochberg (BH) method. To infer the direction of pathway-level alterations, we computed the mean log fold change (logFC) of metabolites associated with each pathway between the normal and pre-pregnant obese groups. A higher logFC in the obese group indicated pathway upregulation, whereas a lower logFC suggested downregulation.

### Ethnicity Association Analysis

To explore ethnicity-related differences in the relationship between metabolites and BMI, we investigated the correlation between metabolite features selected by the ensemble model and clinical data, including maternal ethnicity, paternal ethnicity, and BMI. The Hmisc R package^32^ computed the Spearman correlation coefficients between clinical and metabolomic data. The metabolites that exhibited significant Spearman correlation coefficients for maternal ethnicity were identified as ethnicity-associated metabolites. Significant pathways (p<0.05) were selected for heatmap visualization.

## Results

### Summary of the multi-ethnic cord blood cohort from Hawaii

The multi-ethnic cohort has a total of 87 patients who went through an elective C-section at Kapiolani Women and Children’s Hospital, including 40 pre-pregnant obese mothers and 47 normal-weight mothers before pregnancy (**Table 1**). We elected to investigate C-section due to two considerations: (1) the potential confounding of the laboring process, which would lead to gene expression and subsequent metabolite changes; (2) the fasting states of the women minimize the confounding from the diet. Participants are from three main ethnic groups: Caucasians (N=19), Asians (N=34), and Native Hawaiian/Pacific Islanders or NHPI (N=34). We used stringent matching criteria, such as maternal age, to minimize the confounding effect from these parameters. Indeed, the average ages are similar between the two groups as the study had designed, with 31.25 years in the Control group and 31.38 years in the case group (**Table 1**). As expected, the average BMI values for the control and pre-pregnant obese groups were significantly different, at 21.68kg/m^2^ and 33.88kg/m^2^, respectively. Parity and Gravidity information were also documented and considered in the initial data analysis.

**Table 1.** Cohort Demographics. Demographic characteristics of the cohort, including BMI, maternal and paternal ages, gravidity, parity, and parental ethnicity. Continuous variables (BMI, maternal age, and paternal age) were compared between the control and case groups using t-tests, while categorical variables were compared using chi-square tests.

| Parameter (std) | Control (n=47) | Pre-pregnant obese (n=40) | P-value |
| --- | --- | --- | --- |
| Maternal Age (yr.) | 31.26 (5.69) | 31.38 (4.80) | 0.91 |
| Paternal Age (yr.) | 33.81 (7.20) | 33.4 (6.36) | 0.78 |
| Parity |  |  | 0.0026 |
| 0 | 8 | 4 |  |
| 1 | 30 | 9 |  |
| 2 | 8 | 12 |  |
| 3 and above | 1 | 15 |  |
| Gravidity |  |  | 0.011 |
| 0 | 0 | 0 |  |
| 1 | 7 | 3 |  |
| 2 | 22 | 6 |  |
| 3 and above | 18 | 31 |  |
| Pre-pregnancy BMI (kg/m<sup>2</sup>) | 21.68 (2.04) | 33.88 (4.55) | 2.16 E-21 |
| Maternal Ethnicity |  |  | 0.000851 |
| Caucasian | 12 | 7 |  |
| Asian | 25 | 9 |  |
| NHPI | 10 | 24 |  |
| Paternal Ethnicity |  |  | 0.00231 |
| Caucasian | 13 | 5 |  |
| Asian | 23 | 11 |  |
| NHPI | 11 | 24 |  |

A total of 185 annotated metabolites were selected after the preprocessing step, which included imputation, batch effect removal and normalization (see **Methods**). Besides BMI, other potential confounding variables were assessed using a source of variation (SOV) test (**Supplementary Figure 1**). As expected, BMI is the largest contributor to the variation among the metabolomics data. Other variables with F-statistics >1 (random error) are considered as potential confounders for downstream analysis: maternal and paternal ethnicity, parity, and gravidity. The batch effect is minimal, confirming effective batch correction priorly using ComBat.

### The ensemble regression model significantly outperforms the linear regression model

We constructed an ensemble model consisting of two layers of machine learning models, with three base learners, random forest, XGBoost, and Elastic-net in the first layer and a meta-learner to optimize the performance in the second layer (**Figure 1A**). Given previously reported confounding effects^9^, we calibrated the ensemble model by adjusting the metabolomic data with clinical confounders from SOV analysis above, including maternal and paternal ethnicity, parity, and gravidity, to ensure statistical robustness. The two-layer ensemble model significantly outperformed the baseline linear regression model in 50 iteration tests, with an average RMSE of 4.88 vs. 10.33 (P<0.05), and PCC of 0.67 vs. 0.24 (P<0.05), as compared to the linear regression (**Figure 1B)**. **Figure 1C** shows Shapley’s values (SHAP) of the relative contributions of the top 20 metabolites and clinical confounders to the ensemble model, with the top 3 metabolites being 1,5-anhydrosorbitol (SHAP=1.416), galactonic acid (SHAP=0.818), and glutaconylcarnitine (SHAP=0.712). The importance scores of these metabolites are contributed heterogeneously by the model types in the ensemble. The details of the top 20 metabolites contributing to the ensemble model are listed in **Table 2** with their SHAP values, including 4 fatty acyls, 6 glycerophospholipids, 4 carboxylic acids and derivatives, and 2 organooxygen compounds. Notably, among the demographic variables, parity is ranked first (SHAP score = 1.628) and gravidity third (SHAP score = 0.863) for their importance scores, as indicated in the SOV analysis. Maternal ethnicity NHPI also appears in the top list (SHAP score = 0.299), highlighting its significance in association with the BMI (**Figure 1C**). Overall, the results here demonstrated that integrating both metabolomic and clinical features within a calibrated ensemble metalearner framework provides interpretable insights into the relative influence of biological and demographic factors contributing to maternal BMI.

**Figure 1.**
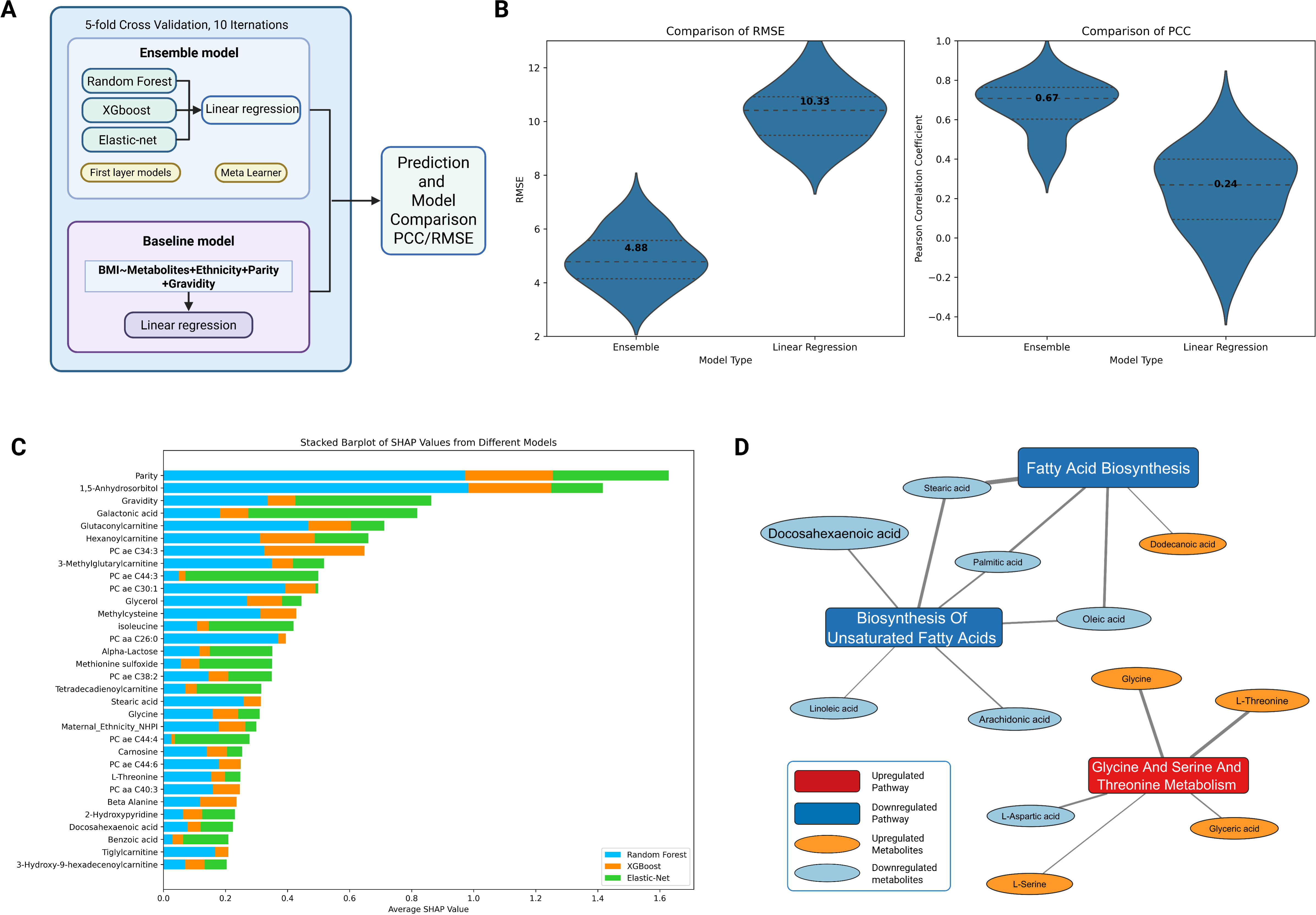
The Ensemble Model Architecture, Performance, Feature Contributions, and Pathway-Level Interpretation. **(A)** The composition of the ensemble model, which consists of two layers. The first layer includes three base learners: Random Forest, XGBoost, and Elastic Net. A meta-learner layer combines predictions from the base learners, assigning weights to each. **(B)** The Performance of the ensemble model in comparison with the baseline linear regression model, using the metrics of RMSE (Root Mean Squared Error) and Pearson correlation coefficient. The model’s performance was evaluated using five-fold cross-validation. **(C)** The top importance of features in the ensemble model is represented by the absolute Shapley value. Feature contributions to the model are color-coded by the base learners: blue (Random Forest), orange (XGBoost), and green (Elastic Net). A higher Shapley value indicates greater feature importance in the model. **(D)** Bipartite graph showing the significantly altered metabolic pathways and their leading edge genes. Pathway nodes and metabolite nodes are sized by their magnitude of change (Log2FC) and colored by the directions of the changes. Edges link each metabolite to its pathway with the width encoding its relative contribution.

**Table 2.** Top 20 Metabolite Features Selected by the Ensemble Model. Key metabolites ranked by their importance to the ensemble model. The SHAP value were derived from the model, * indicates important metabolites overlapped in Schlueter’s study^9^

| ID | Metabolite Biochemical Name | Abbreviation | Class | SHAP |
| --- | --- | --- | --- | --- |
| 1 | 1,5-Anhydrosorbitol * | 1,5-Anhydrosorbitol | monosaccharides | 1.416 |
| 2 | Galactonic acid * | Galactonic acid | Hydroxy acids and derivatives | 0.818 |
| 3 | Glutaconylcarnitine (Mesaconylcarnitine) * | C5:1-DC | Fatty Acyls | 0.712 |
| 4 | Hexanoylcarnitine (Fumarylacarnitine) * | C6(C4:1-DC) | Fatty Acyls | 0.661 |
| 5 | Phosphatidylcholine acyl-alkyl C34:3 * | PC ae C34:3 | Glycerophospholipids | 0.648 |
| 6 | Methylglutarylacarnitine | C5-M-DC | Keto acids and derivatives | 0.518 |
| 7 | Phosphatidylcholine acyl-alkyl C44:3 | PC ae C44:3 | Glycerophospholipids | 0.499 |
| 8 | Phosphatidylcholine acyl-alkyl C30:1 | PC ae C30:1 | Glycerophospholipids | 0.499 |
| 9 | Glycerol | Glycerol | Organooxygen compounds | 0.445 |
| 10 | Methylcysteine | Methylcysteine | Carboxylic acids and derivatives | 0.429 |
| 11 | Isoleucine * | Isoleucine | Carboxylic acids and derivatives | 0.419 |
| 12 | phosphatidylcholine diacyl C26:0 | PC aa C26:0 | Glycerophospholipids | 0.394 |
| 13 | Alpha-Lactose | Alpha-Lactose | Organooxygen compounds | 0.351 |
| 14 | Methionine sulfoxide | Methionine sulfoxide | Carboxylic acids and derivatives | 0.350 |
| 15 | Phosphatidylcholine acyl-alkyl C38:2 | PC ae C38:2 | Glycerophospholipids | 0.350 |
| 16 | Tetradecadienylcarnitine | C14:2 | Fatty Acyls | 0.315 |
| 17 | Stearic acid | Stearic acid | Fatty Acyls | 0.314 |
| 18 | Glycine | Glycine | Carboxylic acids and derivatives | 0.310 |
| 19 | Phosphatidylcholine acyl-alkyl C44:4 | PC ae C44:4 | Glycerophospholipids | 0.278 |
| 20 | Carnosine | Carnosine | Peptidomimetics | 0.254 |

### Pathway analysis shows BMI increase is accompanied by upregulation of amino acid and repression in fatty acid biosynthesis

To investigate the metabolic pathways that may be associated with BMI, we first calculated the pathway deregulation scores by the Pathifier package using 170 metabolites overlapped with the KEGG database. We then associated BMI with each pathway by the Kendall’s tau correlation test with BH adjustment. Three pathways show significance (adjusted p<0.05), including “Biosynthesis of Unsaturated Fatty Acids”, “Fatty Acid Biosynthesis”, and “Glycine, Serine and Threonine Metabolism” (**Supplementary Table 1**). **Figure 1D** shows the bipartite graph of these three pathways and their leading metabolites. The pathways are separated into two modules with opposing deregulation patterns. The first module includes “Biosynthesis Of Unsaturated Fatty Acids” and “Fatty Acid Biosynthesis”, both of which were downregulated with negative logFC comparing obese to normal (logFC = −0.12 and −0.02, respectively) (**Figure 1D, Supplementary Table 1**). The inhibition effect is mainly exhibited by the reduction of stearic acid, oleic acid, and palmitic acid, which are common metabolites between both pathways (**Figure 1D**). The other module is the activated pathway for “Glycine, Serine, and Threonine Metabolism” (logFC = 0.03) associated with an increase in metabolites L-threonine and glycine (**Figure 1D, Supplementary Table 1)**.

### Cord blood L-threonine, 1,5-anhydrosorbitol, glycine and PC(O-44:6) levels are uniquely associated with maternal NHPI ethnicity

Since maternal NHPI ethnicity emerged as a key predictor in the ensemble model (**Figure 1C**), we next examined metabolite associations with maternal ethnicity, with particular attention to NHPI. We examined correlations between the top 20 BMI-associated metabolites and those important clinical variables (**Figure 2A**). Four of these metabolites show both strong correlations and high abundance (above the mean value of all normalized metabolite intensities) with maternal ethnicity: L-threonine, 1,5-anhydroglucitol, glycine, and PC(O-44:6). The changes of average values of these metabolites from normal to obese groups varied across maternal ethnic groups (**Figure 2B-E**). Interestingly, NHPI individuals show the trend of increases for L-threonine, glycine and 1,5-anhydrosorbital as BMI increases, leading to the highest levels in the obese NHPI group. Contrarily, these metabolites are either not changed, or even lower in the obese group than in the controls, among asians and caucasians. L-threonine and glycine are components of the Glycine, Serine, and Threonine metabolism pathway (**Figure 1D**). Furthermore, NHPI mothers showed significantly (P<0.05) lower levels of PC(O-44:6) than the other two groups. PC(O-44:6) is a glycerophospholipid directly related to fatty acid metabolism. It is synthesized through the incorporation of long-chain unsaturated fatty acids into the phosphatidylcholine backbone, and its levels reflect upstream fatty acid biosynthetic activity. Consistent to reduced PC(O-44:6) in cord blood sample born from obese mothers, fatty acid metabolism is repressed, evident by the two enriched BMI-associated pathways: Biosynthesis of Unsaturated Fatty Acids and Fatty Acid Biosynthesis (**Figure 1D**). These findings together imply unique metabolic adaptation among NHPI mothers.

**Figure 2.**
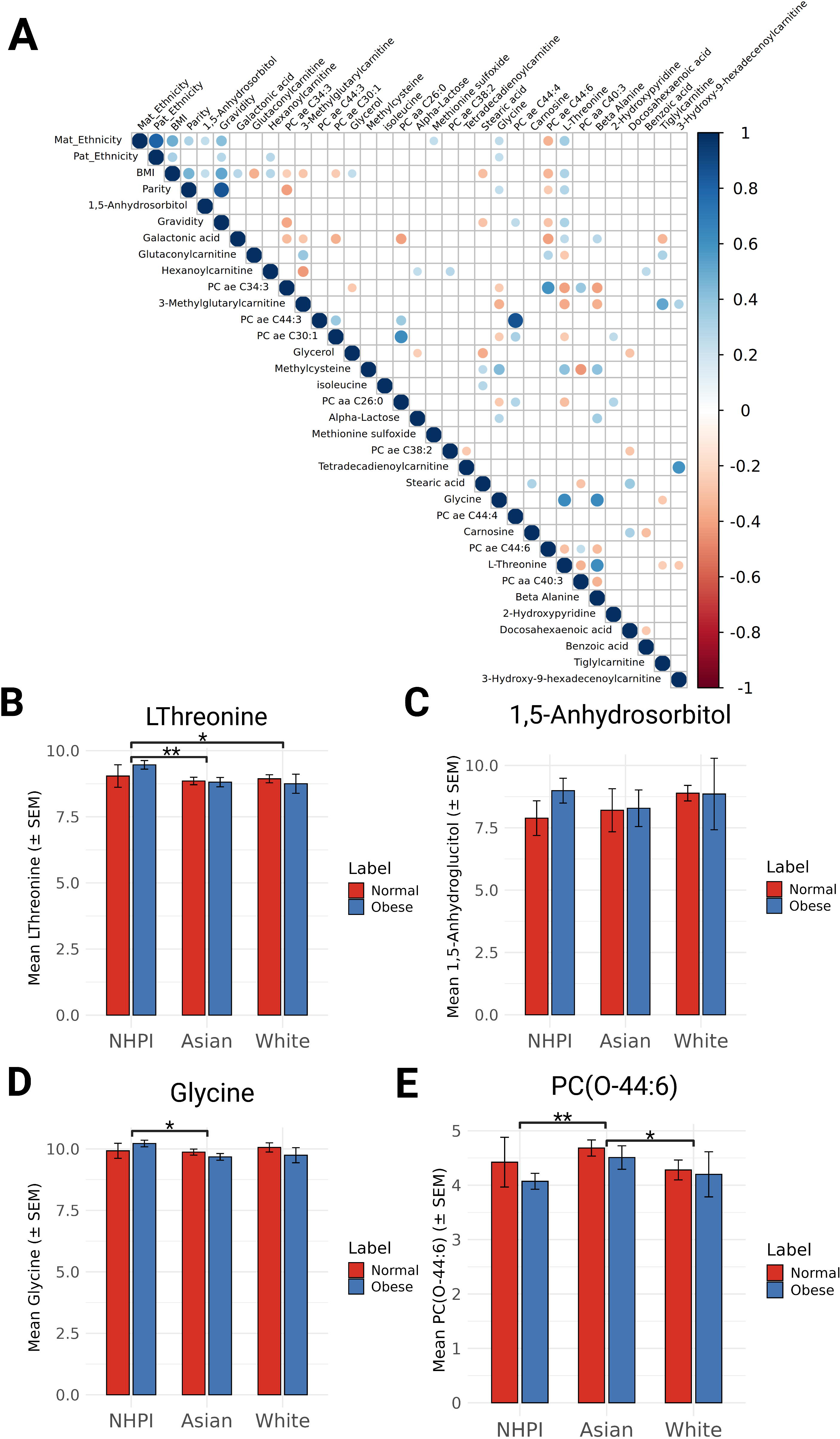
Ethnicity-Stratified Metabolite Differences and Correlations With Clinical Factors. **(A)** Correlation coefficients among important demographical/physiological factors and the metabolomic data. The blue color indicates positive correlations, and the red color indicates negative correlations. **(B-E)** Bar plots showing mean metabolite levels (± SEM) for L-threonine, 1,5-anhydroglucitol, glycine, and PC(O-44:6) across NHPI, Asian, and White maternal ethnic groups, stratified by obesity status (Normal vs. Obese). * p < 0.05; ** p < 0.01. Black lines indicate pair-wise comparison between ethnic groups (normal and obese affiliated cord blood samples combined).

## Discussion

In this study, we aimed to identify more fine-grained associations between cord blood metabolites and maternal obesity. We achieved this from two analytical aspects: (1) we represented obesity as continuous BMI values instead of categories, (2) we used a stacked ensemble machine-learning approach to predict BMI features. Representing obesity as continuous values has many advantages, such as retaining more accurate information of maternal obesity, reducing the risk of biases in metabolite detection, and maintaining statistical power, an issue commonly associated with categorizing continuous variables^33^. The ensemble machine-learning approach has two layers: diverse base learners and a linear regression meta-learner. This design reduces biases and variances inherent to individual base learners, enhances robustness by minimizing overfitting, and improves interpretability by quantifying each learner’s contribution while evaluating both linear and nonlinear relationships between metabolites and BMI.

Among the top 20 important metabolites selected by our ensemble model, Glutaconylcarnitine, Hexanoylcarnitine, Galactonic acid, Isoleucine, Phosphatidylcholine acyl-alkyl C34:3 and 1,5-Anhydrosorbitol overlapped with findings from Schlueter’s previous study (**Table 2**) ^9^. Additionally, the new continuous approach identifies additional metabolites associated with more nuanced variations in BMI that were not revealed before. The new analysis also identified three metabolomic pathways significantly associated with BMI values: Glycine, Serine, and Threonine Metabolism, Fatty Acid Biosynthesis, and Biosynthesis of Unsaturated Fatty Acids. These findings align with and expand upon existing literature. For example, glycine metabolism has been linked to newborn birth weight^34^, consistent with our observation of higher birth weights in infants of obese mothers within the same cohort^35^. This suggests a potential influence of maternal BMI on fetal growth mediated through activation of glycine pathways. On the other hand, metabolites associated with the Biosynthesis of Unsaturated Fatty Acids pathway are downregulated in our cohort, agreeing with findings that higher maternal BMI is associated with reduced fatty acid levels in fetal cord plasma^36^. This downregulation could have implications for fetal growth and development. Finally, the observed downregulation of the Fatty Acid Biosynthesis pathway in the cord blood mirrors reports of decreased lipogenic gene expression in subcutaneous adipose tissue of obese women, suggesting that reduced fatty acid synthesis may serve as a systemic metabolic adaptation to increased adiposity even in newborns of obese mothers^37^.

Our cohort has a significant proportion of Native Hawaiian and Pacific Islander (NHPI) participants, addressing an important gap in health disparities research, particularly given the elevated burden of metabolic diseases such as type 2 diabetes mellitus and obesity observed in this population ^38,39^. Notably, cord blood of babies from NHPI pre-pregnancy obese mothers in our study exhibited distinct metabolic profiles, including elevated L-Threonine, glycine, and 1,5-anhydrosorbitol, but reduced PC(O-44:6), while these metabolites do not show significant changes in asians and caucasians or the opposite changes. Impaired glycine availability may exacerbate systemic inflammation and adiposity, contributing to obesity and related metabolic disorders ^34,40,41^. Similarly, disruptions in 1,5-anhydrosorbitol metabolism can impair glycolysis, increasing susceptibility to obesity^42^. These findings support unique adaptation to maternal obesity in newborns from the NHPI mothers. Future studies to link the ethnicity genetic traits with the metabolite changes in obesity will be of great values.

Several caveats need to be addressed. The relatively small cohort size and limited clinical data constrained our ability to provide more nuanced explanations for observed metabolite aberrations. A larger cord blood cohort with coupled GWAS assay in mothers will help to uncover complex genetic-metabolite links. The clinical EHR data extractable from the patients will also help to filter additional possible confounders ^43–46^. Despite these limitations, this study draws conclusions on critical effects of maternal pre-pregnancy obesity on offspring cord blood metabolites, and provides valuable insights for the modulation of ethnicities, particularly in NHPI populations that are sparsely reported.

## Conclusion

In conclusion, we employed an ensemble machine-learning approach to associate maternal BMI values with newborns’ metabolomic profiles. We observed activation in Glycine, Serine, and Threonine Metabolism, and repression of Fatty Acid Biosynthesis, and Biosynthesis of Unsaturated Fatty Acids in offspring, affected by maternal obesity. In addition, we identified four key metabolites associated with maternal ethnicity: L-Threonine, Glycine, 1,5-Anhydrosorbitol, and PC(O-44:6). These findings enhance our understanding of the metabolic and genetic factors underlying BMI variations, particularly in underrepresented populations.

## Data Availability

The metabolomic raw data files as well as processed data are available at the Metabolomics Workbench repository (https://www.metabolomicsworkbench.org/, study ID ST001114).

https://www.metabolomicsworkbench.org/data/DRCCMetadata.php?Mode=Study&StudyID=ST001114

## Acknowledgments

This study was supported by the National Institutes of Health grants R01 LM012373 and LM012907 from the National Library of Medicine (NLM), and grant R01 HD084633 from the Eunice Kennedy Shriver National Institute of Child Health and Human Development (NICHD), awarded to L.X. Garmire. This research was supported in part by training funding provided by the NIH grant T32 GM141746 and Advanced Proteogenomics of Cancer (T32 CA140044). Through a secondary analysis of a previous study, we thank the previous colleagues of the OBGYN department and institute of biogenesis research (IBR) of University of Hawaii Medical School, in helping collect the cord blood samples.

## Author’s contributions

LG envisioned this project, obtained the funding, supervised the study, and revised the manuscript. LT performed the data analysis, and wrote the initial manuscript. BL modified the analysis, generated the figures, and wrote part of the manuscript. YD advised both LT and BL to perform the analyses, troubleshoot and revised the manuscript. SH helped with the analysis and edited the manuscript. All authors have read the manuscript.

---

**Supplementary Table 1:** Pathway Correlation with BMI. Significant results of the Kendall’s tau correlation test between 29 mapped pathways and continuous BMI values.

| Pathway name | BH adjusted P.val | PDS_value | Correlation | Mean Metabolites logFC |
| --- | --- | --- | --- | --- |
| Glycine, Serine And Threonine Metabolism | 0.01 | 0.52 | 0.19 | 0.03 |
| Biosynthesis Of Unsaturated Fatty Acids | 0.02 | 0.44 | 0.17 | -0.12 |
| Fatty Acid Biosynthesis | 0.03 | 0.55 | 0.16 | -0.02 |

