## Supplementary Figure 1 for "An ensemble method associates prepregnancy BMI and maternal ethnicity with key cord blood metabolomic changes in a multi-ethnic cohort from Hawaii"

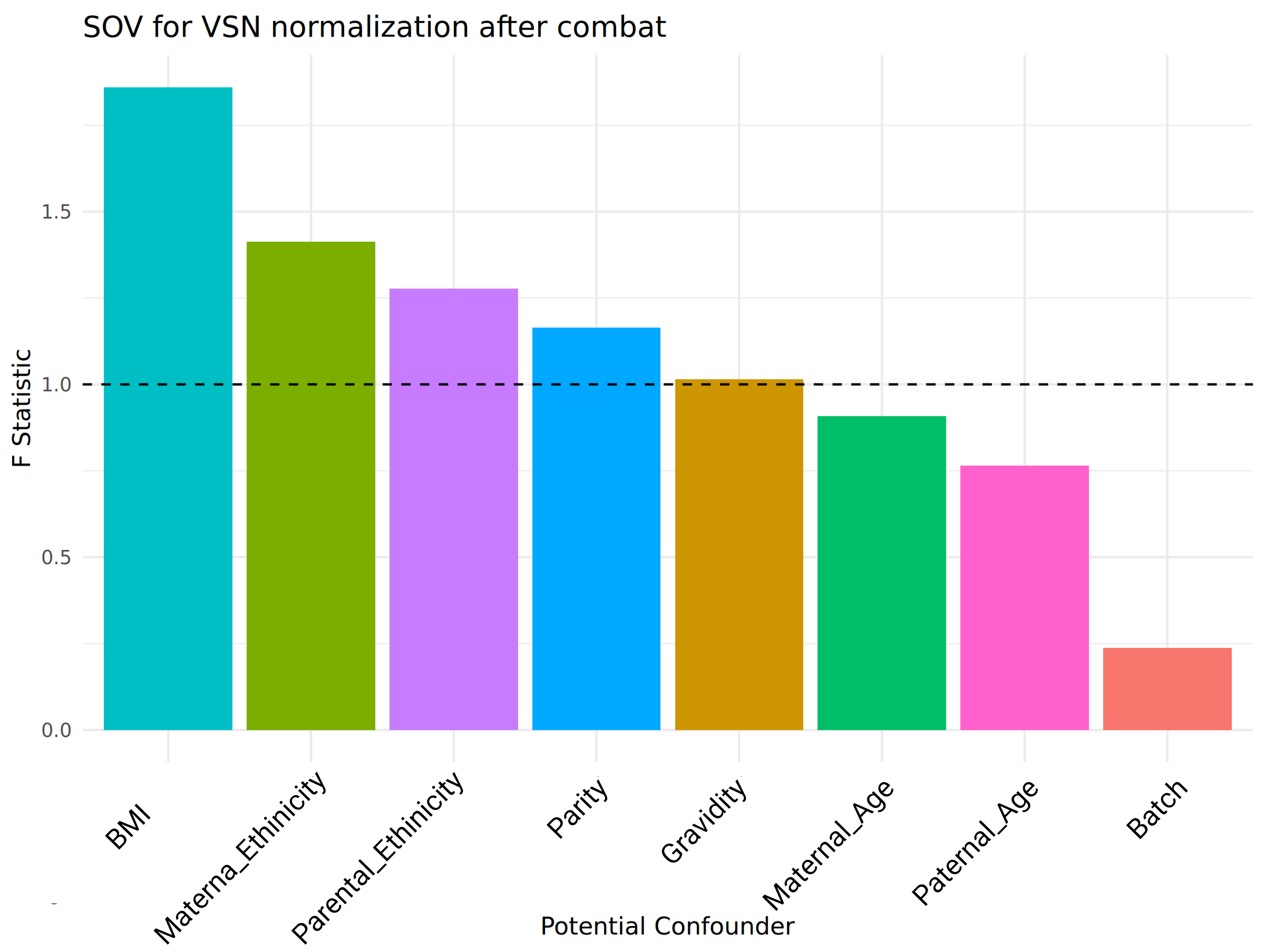

**Supplementary Figure 1. Source of Variance Analysis**.

The significance of each variable was assessed using source of variance analysis per ANOVA, with F-statistics calculated for each variable. The red line represents the threshold for calling confounders (F ratio = 1). Sample grouping (BMI) is the dominant factor to explain the difference among the metabolomics data. Four other factors are identified as confounders: maternal ethnicity, paternal ethnicity, parity, gravidity. Batch effect has been successfully removed, as shown by the figure (F ratio <1).
